# A neural signature of touch aversion and interpersonal problems in Borderline Personality Disorder

**DOI:** 10.1101/2025.01.10.25320260

**Authors:** Jella Voelter, Danilo Postin, Ilona Croy, René Hurlemann, Dirk Scheele

## Abstract

**Introduction:** Patients with borderline personality disorder (BPD) suffer from severe social impairments and interpersonal problems. Social touch can provide comfort and facilitate the maintenance of social bonds, and preliminary evidence indicates a negative evaluation of social touch in patients with BPD. However, the neural mechanisms underlying aberrant touch processing in BPD and its role for social impairments are still unclear.

**Methods:** We recruited 55 BPD patients and 31 healthy controls and used functional magnetic resonance imaging to probe neural responses to slow (i.e. C-tactile (CT)-optimal; affective) and fast (i.e. CT-suboptimal; discriminative) touch before and after four weeks of a residential dialectical behavior therapy (DBT) program. In addition to assessing BPD symptoms and interpersonal problems, we evaluated touch allowance maps and the attitude towards social touch.

**Results:** BPD patients showed a comprehensive negative bias towards social touch before the DBT, evident in a significantly more negative attitude towards and reduced comfort zones of social touch compared to healthy controls. Moreover, reduced comfort zones of social touch were associated with more interpersonal problems. Activation in the posterior insular cortex in response to CT-optimal touch was significantly reduced and correlated with the severity of interpersonal problems in BPD patients. Despite significant improvements in overall BPD symptom load, dysfunctional social touch processing persisted after four weeks of DBT, indicating trait-like disturbances in BPD.

**Conclusions:** An impaired insula-mediated integration of affective and sensory components of touch may constitute a clinically relevant biological signature of the complex interpersonal problems in BPD.

## Introduction

*“[…] Borderline individuals are the psychological equivalent of third-degree burn patient[s]. They simply have, so to speak, no emotional skin. Even the slightest touch or movement can create immense suffering.”* [1, p. 69].

In 1993, Marsha Linehan, creator of the dialectical behavior therapy (DBT), illustrated a problematic connection between touch and affective processing in patients with borderline personality disorder (BPD). Over 30 years later, our understanding of touch perception in this versatile psychiatric disorder remains limited. BPD is a costly psychiatric disorder, highly prevalent in clinical settings and associated with premature death and severe functional and social impairments [2–4]. Unstable interpersonal relationships, affective dysregulation, impulsivity, and identity disturbances represent the core symptoms of BPD [3,5,6]. Interpersonal problems as a key component of BPD manifest in a negative bias towards social cues, impaired interpersonal trust, and hypersensitivity to social exclusion and threat [7]. As such, BPD patients report increased loneliness [8] and impaired global social functioning [9]. Psychotherapy is the treatment of choice for BPD, with DBT and psychodynamic approaches being the most effective [2]. DBT is based on cognitive behavioral therapy and integrates strategies of acceptance and change, with a focus on improving emotional regulation, dysfunctional behaviors, interpersonal problems, and mindfulness [1]. Residential DBT involves group and individual treatment sessions simultaneously [10]. The etiology of BPD is strongly linked to adverse childhood experiences, such as neglect and physical abuse [11] that in interaction with genetic factors lead to altered neural development and increased risk for BPD [2]. Meta-analytic evidence indicates amygdala hyperreactivity to emotional stimuli as well as altered reactivity in the prefrontal and insular cortex during emotional processing in patients with BPD [12,13]. Single studies also report hyperreactivity to negative stimuli in striatal areas [14,15] and reduced amygdala habituation to repeated negative stimuli [16]. However, it is still unclear whether amygdala hyperreactivity and altered reactivity in the insular cortex and striatum also affect the processing of positive social stimuli and how this may contribute to social impairments in BPD.

Interpersonal touch is crucial for social communication and maintaining social bonds across cultures worldwide [17]. Touch is a powerful tool to communicate emotions [18] and comfort conspecifics [19]; it can further decrease stress responses [20], and alleviate pain [21]. Interpersonal touch involves the transmission of both sensory and emotional information through different mechanoreceptor afferents. Unmyelinated C-tactile (CT) afferents in the non-glabrous skin provide information about the emotional-motivational properties of touch, i.e., affective touch. These fibers respond best to slow, gentle stroking at a speed of 1-10 cm/s and project to the posterior insula cortex, a key region for interoceptive processing [22,23]. Affective components of touch are additionally processed in other socioemotional brain networks, including striatal reward structures [24].

While an altered processing of pain in BPD patients is well established [25], research on touch processing remains scarce. Preliminary evidence indicates perceptual changes in touch processing, including descriptions of touch as rougher, firmer, and less intense, as well as heightened thermal and somatosensitive thresholds in patients with BPD [26,27]. Furthermore, BPD patients report a more negative attitude towards social touch (e.g., less liking and less frequency of social touch; [28]). However, the neural underpinnings of altered touch processing and its role for interpersonal problems in BPD patients have not been tested yet. More severe childhood maltreatment correlates with reduced comfort of and heightened responses to fast (i.e. CT-suboptimal) touch in the posterior insula and somatosensory cortex [29]. By contrasts, major depressive disorder (MDD) patients also rated CT-suboptimal touch as less comforting, but they exhibited reduced striatal responses to touch compared to healthy controls (HC) [30]. Given the high prevalence of childhood maltreatment [11] and MDD comorbidity [2], impaired touch processing in BPD patients could be related to altered sensory processing in the insula cortex [26,27], amygdala-based hypervigilance to social threats [16,31], or a negative striatal bias towards positive social cues [24,32]. Interestingly, DBT treatment effects were observed in the amygdala and anterior insula cortex in response to the evaluation and reappraisal of negative stimuli [33,34]. A central part of DBT is training mindfulness. Through DBT, BPD patients learn to be aware of their experiences in a non-judgmental way, which involves exercises engaging the sense of touch. Another key focus of DBT is increasing interpersonal effectiveness by teaching interpersonal problem-solving strategies through effective assertiveness, behavioral reinforcement, and empathy or validation skills [35]. Given the efficacy of DBT in improving psychosocial functioning [10,36], it is conceivable that DBT affects the perception of interpersonal touch by enhancing mindfulness and improving emotion regulation strategies.

Thus, in the present study, we used functional magnetic resonance imaging (fMRI) to decipher neural responses to slow (i.e. CT-optimal) and fast (i.e. CT-suboptimal) social and non-social touch in BPD patients before and after four weeks of a residential DBT program. To further characterize BPD patients and to control for naturally occurring changes in the outcome measures, we included a control group involving HC that did not receive an intervention. The first of our pre-registered hypotheses stated that BPD patients would exhibit a more negative attitude towards and altered processing of social touch compared to HC at pre-measurement (i.e. before the DBT). More specifically, given the consistently reported hyperactivity of the amygdala to socially threatening stimuli in BPD [3,16], we expected that BPD patients compared to HC would display increased activity and reduced habituation in the amygdala in response to touch. We further anticipated altered activity in the insular cortex, a key region involved in the processing of affective touch [22,23]. This is supported by previous findings of aberrant insular processing during touch in individuals with more severe childhood maltreatment [29], which is common in BPD. Finally, we hypothesized decreased activity and enhanced habituation in striatal areas in response to touch, which may reflect reduced hedonic value of social touch as observed in patients with MDD [30]. Additionally, we explored associations with interpersonal problems, symptom severity, and childhood maltreatment. Furthermore, we hypothesized that BPD-associated impairments would be reduced after DBT treatment and that these treatment effects would be more pronounced in patients with stronger symptom reduction.

## Methods

### Study Design and Participants

The study design was registered at ClinicalTrials.gov (NCT04770038), and the analysis plan was pre-registered before conducting any analyses (https://osf.io/zn5c7). A total of 55 BPD patients on a waiting list for DBT were recruited at the Karl-Jaspers-Klinik in Bad Zwischenahn, Germany. Trained clinical psychologists at the DBT outpatient clinic confirmed the BPD diagnoses through interviews as part of their clinical work. Data from 55 BPD patients were compared to 31 healthy participants without any psychiatric illness (see **Supplementary Information (SI)** for comorbidities and medication of the patients). General exclusion criteria were age under 18 or over 65 years, MRI contraindication, scars on a predefined area of 20 cm of their shins, acute suicidality, any lifetime psychotic disorders, current substance dependence, a history of traumatic brain injuries, or other neurological illnesses. The presence of psychiatric disorders or current or past psychiatric inpatient treatment resulted in exclusion from study participation in the HC group. Longitudinal analysis included 37 BPD patients, as 18 BPD patients either did not receive residential treatment during the study period or left treatment early, and 31 healthy controls (HC; see **Supplementary** Fig. 1). The sample size was based on an a priori power-analysis (see **SI Power Analysis**). For details on demographic and clinical characteristics, see **Supplementary Tab. 1-3**.

### Residential DBT program

BPD patients register themselves at the DBT outpatient clinic and are invited to a first interview where a detailed assessment, discussion of life circumstances, and evaluation of treatment options takes place. This is followed by a pre-inpatient group, where treatment goals are discussed. The residential DBT program at the Karl-Jaspers-Klinik is structured into three modules, each lasting four weeks, with a varying outpatient practice phase in between. During these phases, patients are advised to apply the skills they have learned during therapy to their everyday lives. The first DBT module at the Karl-Jaspers-Klinik prioritizes the reduction of suicidal and parasuicidal behaviors, enhancing stress tolerance, managing cravings, and addressing dissociations. This module can function as a standalone treatment unit. Following this, the second module focuses on understanding and regulating emotions. The last module provides the opportunity to enhance interpersonal skills, foster self-esteem, and enhance overall quality of life. The treatment sessions include both group and individual therapy. During the group sessions, patients from all three modules participate together. The group sessions focus on psychoeducation and skills training, as outlined in the DBT skills training manual [35]. The skills training forms the core of the DBT program. Patients learn strategies to effectively distract and calm themselves during high-stress situations, how to address interpersonal problems, and how to better perceive and regulate emotions through mindfulness. On weekdays, patients typically return home to engage in a stress trial in their usual environment. BPD patients were measured before (pre-measurement) and after one module (i.e., four weeks) of the residential DBT program (post-measurement). The HC underwent the same measurements but with no intervention between pre- and post-measurement.

### Psychological and Clinical Assessments

The Borderline Symptom List-23 (BSL-23) [37] was utilized to evaluate the severity of BPD symptoms and the Childhood Trauma Questionnaire (CTQ) [38] was used to assess childhood trauma. The Inventory of Interpersonal Problems (IIP) was administered to measure severity of interpersonal problems [39] and the Social Touch Questionnaire (STQ) [40] was applied to assess the attitude towards social touch. For further questionnaires, see **SI Psychological and Clinical Assessments**. BPD patients were subdivided into treatment responders and non-responders by calculating the Reliable Change Index (RC) using the BSL-23 as outcome variable. The RC determines whether an individual difference between two measurements (pre/post) reflects more than a random measurement error by considering the reliability of the measurement instrument [41]. RCs of less than 1.96 were classified as no treatment response, RCs equal or above 1.96 were classified as a treatment response.

### Comfort Zones of Social Touch

Comfort zones of social touch were assessed with a computerized task [42]. Participants indicated on a human silhouette representing their own body, where a specific social network member (e.g., their mother) would be allowed to touch them in everyday situations (see **Supplementary** Fig. 2). The human silhouette was divided into the front and back areas and was presented for a total of nine different social network members (friend (f/m), stranger (f/m), partner, brother, sister, mother, father). Participants had the option to skip social network members that did not exist in their social network, e.g. in cases where they had no siblings. To better quantify comfort zones of social touch, a Touchability Index (TI) was calculated. The TI represents the proportion of colored pixels within the body mask ranging from 0 (0% pixel filled out) to 1 (100% pixel filled out).

### fMRI Touch Task

Participants underwent an adapted version of an fMRI touch paradigm [29]. Conditions consisted of four types of touch trials with combinations of CT-optimal (∼ 5 cm/s) and CT-suboptimal touch (∼ 20 cm/s), as well as social (administered by hand) and non-social (administered with a brush) touch and a control (no touch) condition (**Fig. 1A**). An experimenter, who was unknown and not visible to the participant to control for sex differences and familiarity, administered the touch for four seconds across 20 cm of both shins. Types and duration of touch were signaled via tones to the experimenter wearing headphones and the experimenter was trained to apply the tactile stimuli with a consistent speed and pressure. During the experiment, participants rated the comfort of each trial on a visual analogue scale (VAS) from 0 (not at all comforting) to 10 (very comforting), including no touch trials. Each of the five conditions (CT-optimal social touch, CT-optimal non-social touch, CT-suboptimal social touch, CT-suboptimal non-social touch, no touch) compromised 12 trials, resulting in a total of 60 trials and a measurement time of ∼16 minutes. The task was divided into two runs of 30 trials each, with a 30-second break in between. Each rating lasted six seconds.

**Fig. 1.**
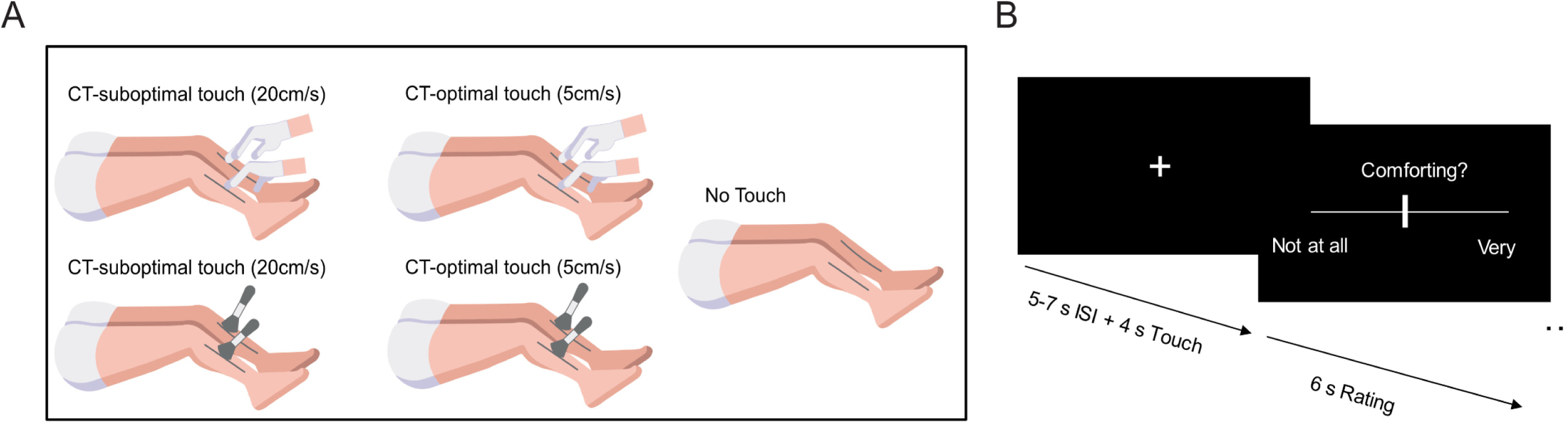
Conditions of the fMRI touch task (A). Touches were applied to the shins for four seconds. The touch stimuli differed in velocity (CT-optimal speed: 5 cm/s and CT-suboptimal speed: 20 cm/s), and sociality (social touch administered with hand and non-social touch administered with brush). Task design (B). After each trial, including the no touch trial, participants rated the comfort of the trial on a VAS. The ratings lasted six seconds, and the interstimulus interval (ISI) duration was randomized between five and seven seconds.

### MRI Data Acquisition and Preprocessing

MRI data were acquired using a 3T Siemens Prisma MRI scanner (Siemens AG, Erlangen, Germany) with a 64-channel head coil. High-resolution anatomical images were measured with a T1-weighted 3D MP-RAGE sequence. A T2*-weighted echoplanar multiband sequence with a multiband acceleration factor of 4 [43] was used to measure neural responses to touch. fMRI data were preprocessed using the standardized pipeline *fMRIPrep* 20.2.1 [44].

### fMRI Data Analysis

The fMRI analysis in SPM12 involved a two-level approach based on the General Linear Model. Treatment effects were investigated by calculating differences between pre- and post-measurement data in BPD patients and HC on the first level. Second-level statistical inference included two-sample and one-sample t-tests. Sex, age, Body Mass Index (BMI), DBT module, and prescan inner tension (assessed with a VAS) were included as covariates. The analysis was carried out using a single Region-of-Interest (ROI) mask comprising the anatomically defined amygdala, the insular cortex, and striatal areas. To test whether group effects (BPD vs. HC) on neural responses to touch vs. no touch and CT-suboptimal vs. CT-optimal touch were moderated by childhood trauma or BPD symptom severity, t-tests were calculated with an additional interaction term (e.g., group x CTQ scores). Likewise, we tested whether changes in symptom severity (BSL scores pre minus post) and treatment response (responder vs. non-responders) moderated differences between the pre- and post-measurement for the contrast touch vs. no touch. Significance was assessed at peak level with *p* < 0.05, family-wise error (FWE) corrected.

### Behavioral Data Analysis

Two-sample t-tests were used to compare questionnaire data and the TI between groups, and paired-t tests were applied to investigate within-group effects. Mixed-Design ANOVAs were performed to compare differences in comfort ratings of touch. Multilevel modeling with repeated measures was performed to investigate whether group differences (BPD vs. HC) in TI were specific to social network members and to investigate treatment-related effects. Touch allowance maps were compared between groups by two proportion z-tests and within groups by McNemars tests, False Discovery Rate (FDR) corrected [45]. Additional mixed-design ANOVAs were conducted to compare changes in STQ scores, the TI, and comfort ratings of touch in BPD patients to changes in the HC group. Moderation analyses were carried out to assess whether childhood trauma or BPD symptom severity moderated group differences (BPD vs. HC) in STQ scores, the TI, and comfort ratings of touch. Likewise, we examined whether treatment effects differed between DBT responders and non-responders and were moderated by changes in BSL scores. For all analyses, sex, age, and BMI were included as covariates. Therapy modules were included as a covariate for analyses investigating within-subject effects of the patient group. Exploratory correlational analyses between CTQ scores, BSL scores, IIP scores, and the significant parameter estimates of the fMRI analysis, as well as the TI, were carried out using Pearson’s correlation coefficient. Statistical significance was assessed at *p* < 0.05 (two-tailed tests for undirected hypotheses and one-tailed tests for directed hypotheses). Post-hoc comparisons were corrected for multiple comparisons using a Bonferroni-Holm correction (*p*_cor_). For details of the fMRI and behavioral analyses see Supplementary Methods.

## Results

Before the treatment, BPD patients exhibited a high symptom load (M±SD: 1.98±0.87) that was significantly different from the ratings of HC (0.20±0.16; *t*_(60.47)_=14.73, *p*<0.0001*, d*=2.53; see **Fig. 2A**). Likewise, BPD patients reported significantly more severe childhood maltreatment (CTQ score: 65.00±21.00; see **Fig. 2B**), a higher degree of interpersonal problems (IIP score: 2.16±0.47; see **Fig. 2C**), and more pronounced social touch aversion (STQ score: 45.04±10.88; see **Fig. 2D**) than HC (CTQ score: 35.06±10.03; *t*_(82.24)_=8.90*, p*<0.0001*, d*=1.67; IIP score: 0.98±0.40; *t*_(84)_=11.76*, p*<0.0001*, d*=2.64; STQ score: 27.48±8.47; *t*_(82)_=7.71*, p*<0.0001*, d*=1.74). Furthermore, interpersonal problems correlated positively with social touch aversion across both groups (*r*_(82)_=0.60, *p*<0.0001).

**Fig 2.**
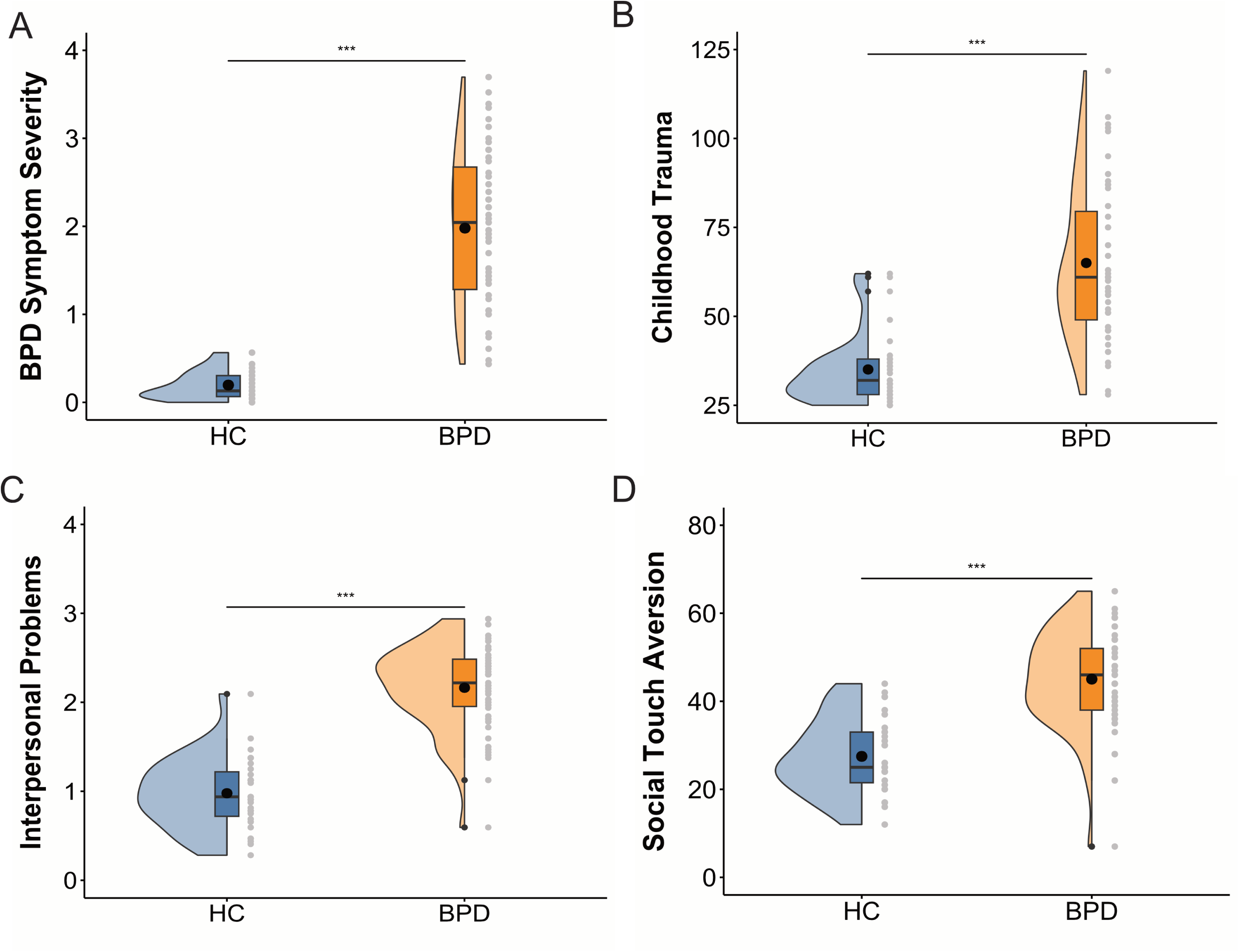
At baseline, BPD patients (n=55) reported significantly higher symptom severity (BSL-23 scores, A), childhood trauma experiences (CTQ scores, B), interpersonal problems (IIP scores, C) and social touch aversion (STQ scores, D) compared to HC (n=31). In the boxplot, the line dividing the box and the black dot represent the median and mean of the data. The ends represent the upper/lower quartiles and the extreme lines represent the highest and lowest values excluding outliers. Abbreviations: BPD, borderline personality disorder; HC, healthy controls; \*\*\**p*<0.001.

BPD patients reported a significantly reduced TI compared to HC *(t*_(80)_=-6.06*, p* <0.0001*, d* =-1.38) indicating that they considered touch to be less acceptable across social network members (see **Fig. 3A**). Furthermore, the TI correlated negatively with interpersonal problems (IIP scores) in BPD patients (*r*_(49)_=-0.32, *p*=0.02) and across both groups (*r*_(80)_=-0.54, *p*<0.0001). A multilevel model with the TI as dependent variable revealed significant main effects of social network member (*F* 149.25*, p* <0.0001, η^2^=0.68) and group (*F* 39.05*, p* <0.0001, η^2^=0.34), and a significant interaction effect of group and social network member (*F* =5.39*, p* <0.0001, η^2^=0.07, see **Supplementary Tab. 4**). Post-hoc pairwise comparisons showed significant differences between BPD patients and HC for all social network members except the partner and female and male strangers (see **Supplementary Tab. 5**). Statistical group comparison of the touch allowance maps by two-proportion z-tests confirmed significantly fewer comfort zones of social touch for BPD patients and illustrate that group differences were not limited to specific body regions and were most pronounced for female friends and family members/parents (see **Fig. 3B** and **3C**).

**Fig 3.**
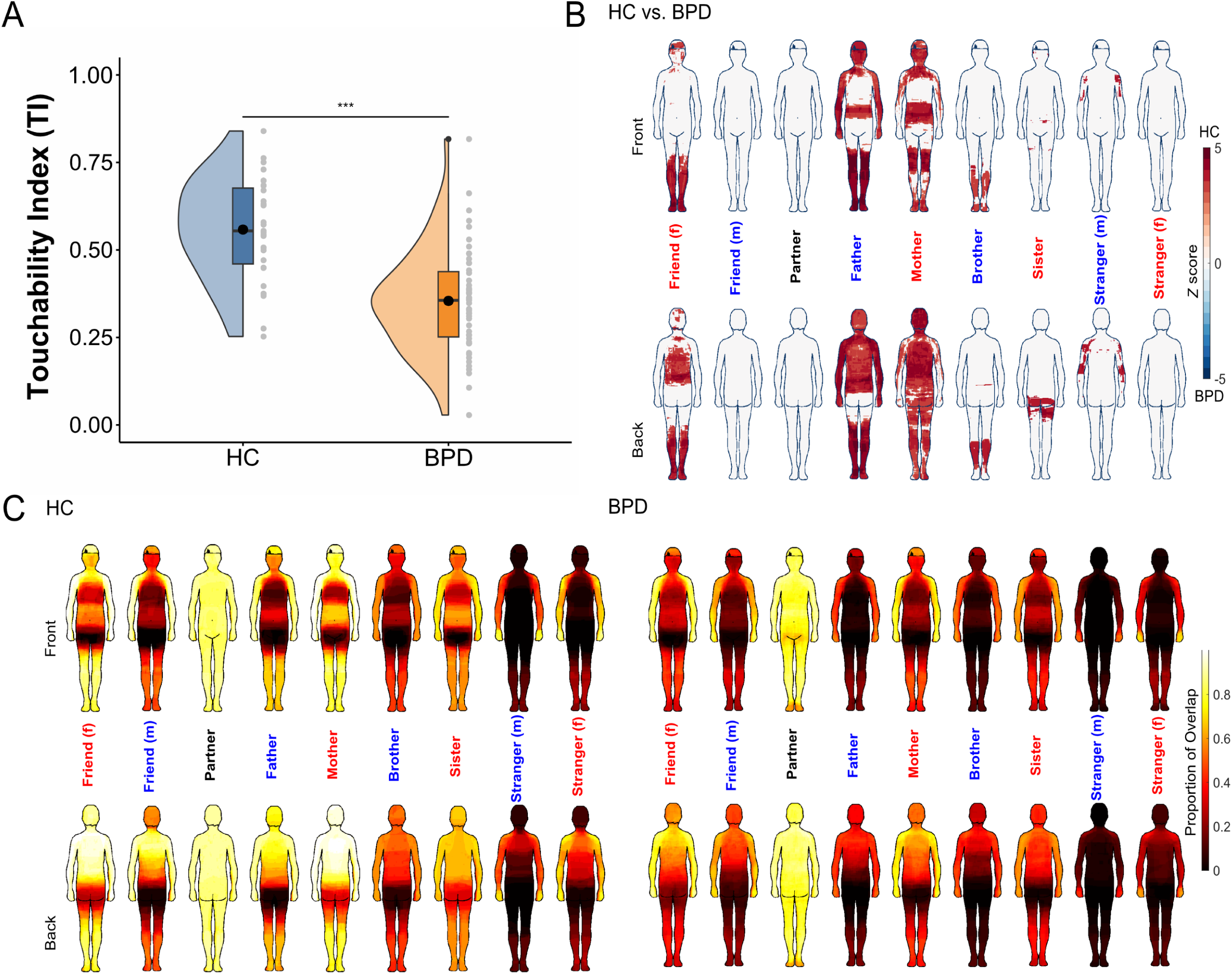
BPD patients (n=51) showed a significantly lower TI across social network members than HC (n=31), i.e., fewer comfort zones of social touch (A). Comparison of touch allowance maps between HC and BPD patients via two-proportion z-tests (B). Reduced touch allowance in BPD patients (in red) were not limited to specific body regions and most pronounced for female friends and family members/parents. Touch allowance maps for HC and patients with BPD (C). The coloring represents the proportion of the sample reporting a body part as an acceptable touch zone for the respective social network member. In the boxplot, the line dividing the box and the black dot represent the median and mean of the data. The ends represent the upper/lower quartiles and the extreme lines represent the highest and lowest values excluding outliers. Abbreviations: BPD, borderline personality disorder; HC, healthy controls; \*\*\**p*<0.001.

In accordance with our hypothesis, BPD patients exhibited significantly altered responses to CT-suboptimal touch relative to CT-optimal touch in the right posterior insula cortex (MNI: 40, -4, 2; *t*_(75)_=4.52*; p* _FWE_=0.037). The extracted parameter estimates of the significant peak voxel revealed significantly reduced insular activation to CT-optimal touch in BPD patients compared to HC (*t*_(79)_=2.50*, p*_(cor)_*=*0.04*, d*=0.54; see **Fig. 4A**). The parameter estimates for CT-optimal touch negatively correlated with interpersonal problems across both groups (*r*_(79)_=-0.39, *p*<0.001) and in BPD patients (*r*_(48)_=-0.34, *p*=0.015, see **Fig. 4B**), but not in HC (*r*_(29)_=-0.28, *p*=0.13). A mediation analysis using group (HC, BPD) as a predictor, parameter estimates of the significant peak voxel as a mediator, and interpersonal problems as the outcome measure further supported this result by revealing a significant partial mediation effect (*B*=0.08, *p*=0.02) of the posterior insula activity on the relation between group and interpersonal problems. There were no significant habituation effects, or group differences for the amygdala and striatal areas or when comparing touch versus no touch and social versus non-social touch.

**Fig. 4:**
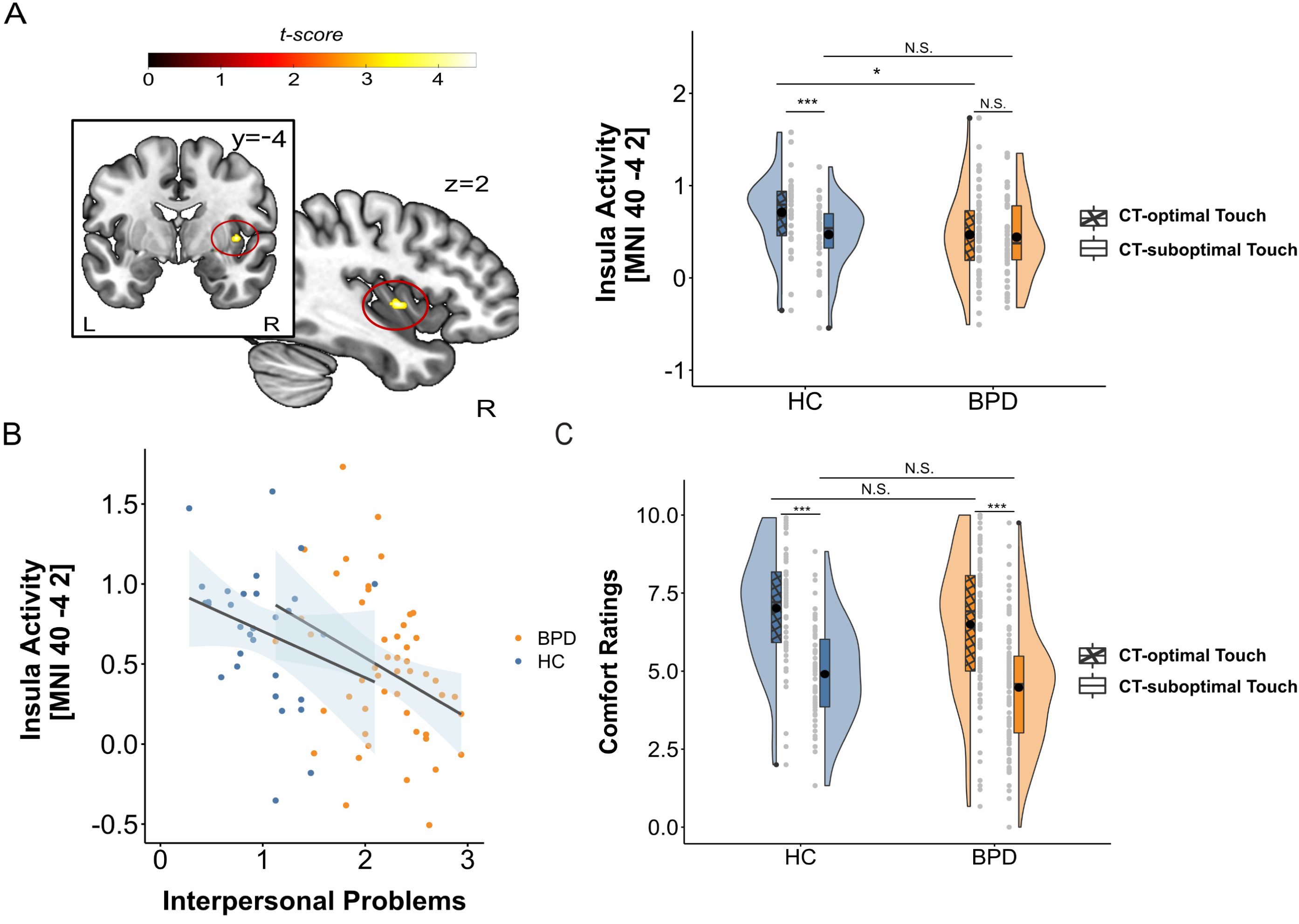
At baseline, patients with BPD (n=50) compared to HC (n=31) showed significantly reduced responses to CT-optimal touch in the right posterior insular cortex (A; MNI: 40, -4, 2). Insula activity for CT-optimal touch correlated negatively with severity of interpersonal problems in BPD patients and across both groups (B). Both HC (n=31) and patients with BPD (n=51) rated CT-optimal touch as more comforting than CT-suboptimal touch (C). In the boxplot, the line dividing the box and the black dot represent the median and mean of the data. The ends represent the upper/lower quartiles and the extreme lines represent the highest and lowest values excluding outliers. Abbreviations: BPD, borderline personality disorder; HC, healthy controls; MNI, Montreal Neurological Institute; \**p*<0.05; \*\*\**p*<0.001. A Bonferroni-Holm correction was applied to adjust for multiple comparisons.

The mixed-design ANOVA with group as between-subject factor (HC, BPD) and velocity (CT-optimal, CT-suboptimal) and sociality (social, non-social) as within-subject factors revealed no group effect nor an interaction effect with group (all *p*-values > 0.16) but a significant effect of velocity *(F*_(1,80)_=78.00*, p*<0.0001, η ^2^*=*0.22). As expected, CT-optimal touch (HC: 7.01 ± 1.80, BPD: 6.50 ± 2.17) was rated more comforting than CT-suboptimal touch (HC: 4.00 ± 1.61, BPD: 4.47 ± 1.90) in both groups (all *p*- values <0.0001) (see **Fig. 4C**). There was no significant difference between social and non-social touch in BPD patients or HC (*F*_(1,80)_=0.001*, p* =0.97, η ^2^*<.*01). A significant interaction was found between sociality and velocity (*F* =24.00*, p<.0001,* η ^2^=.01). Data inspection suggested that CT-suboptimal touch was rated as more comforting when applied by brush (4.87 ± 1.61) than by hand (4.40 ± 1.96), while CT-optimal touch was rated as more comforting when applied by hand (6.89 ± 2.10) than by brush (6.50 ± 1.99). There were no significant moderation effects of symptom severity (BSL-23 scores) or childhood trauma (CTQ scores) on baseline groups differences.

## Longitudinal Results

After four weeks of a residential DBT program, BPD symptom severity showed significant improvement (*t*_(36)_=6.14*, p*<0.0001*, d*=1.06), with a decrease from a high (2.10 ± 0.82) to a moderate symptom load (1.22 ± 0.83; see **Fig. 5A**). Out of 37 patients with longitudinal data, 15 (41%) were classified as responders and 22 (59%) as non-responders. Interpersonal problems decreased after treatment in BPD patients (*t*_(36)_=2.83, *p*=0.007, *d*=0.43, see **Fig. 5B**). However, there were no significant changes in the total TI *(t*_(36)_=1.21*, p* =0.88*, d*=-0.12; see **Fig. 5C**) or social touch aversion *(t*_(36)_=0.25*, p* =0.40*, d* =0.03; see Fig. 5D) after treatment. A statistical comparison using McNemars tests confirmed no significant differences between the pre- and post-treatment touch allowance maps. Likewise, a multilevel model with repeated measures, including the nine social network members, group, and time (pre, post) as categorical fixed effect factors, showed no significant effect of time (with pre as reference category) or an interaction effect of time and group on the total TI (all p-values > 0.18, see **Supplementary Tab. 6**).

**Fig. 5.**
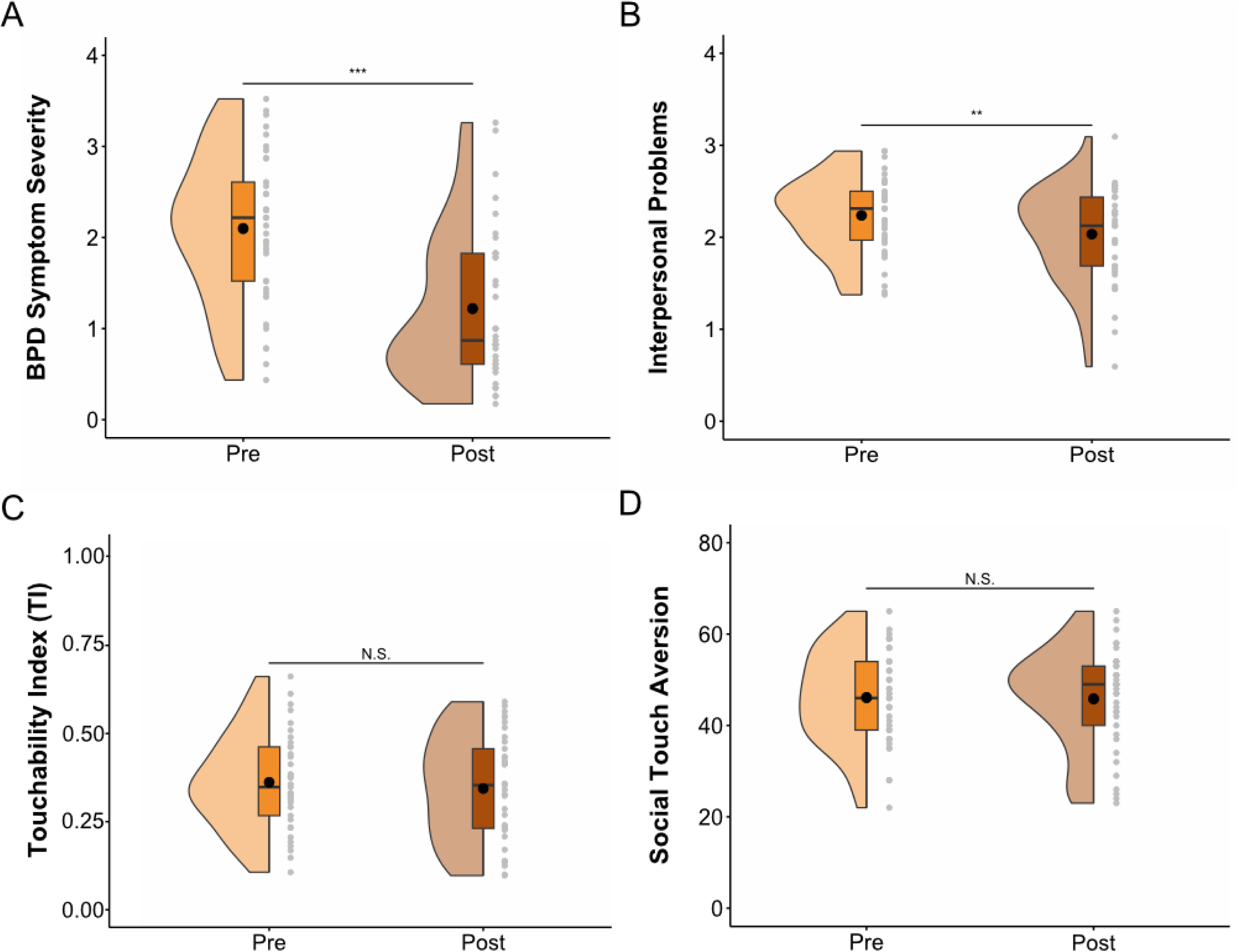
Four weeks of DBT were associated with a significant decrease in BPD symptom severity (BSL- 23 scores, A) and interpersonal problems (IIP scores, B) in BPD patients (n=37). However, there were no significant changes in comfort zones of social touch (total TI, C), and social touch aversion (STQ scores, D). In the boxplot, the line dividing the box and the black dot represent the median and mean of the data. The ends represent the upper/lower quartiles, and the extreme lines represent the highest and lowest values, excluding outliers. Abbreviations: BPD, borderline personality disorder; \*\**p*<0.01; \*\*\**p*<0.001.

Interestingly, a comparison of pre- and post-treatment fMRI data of BPD patients revealed a significant change in the response to CT-optimal vs. CT-suboptimal touch in the right anterior (MNI: 50, 10, -2; *t*_(26)_=5.34*; p*_FWE_=0.036) and posterior insula cortex (MNI: 42, -16, -4; *t*_(26)_=5.58*; p*_FWE_=0.021). However, these changes in BPD patients were not significantly different from HC, suggesting that these changes may be time-dependent. Furthermore, we detected no significant treatment-related changes for other contrasts or regions, or any effects on amygdala and striatum habituation. There was no significant treatment-related effect on comfort ratings of touch stimuli (all p-values > 0.55).

Moderation analyses revealed that treatment-associated differences (pre vs. post) for social touch aversion (STQ total score), comfort zones of social touch (total TI), comfort ratings for touch stimuli, and neural responses to touch vs. no touch were not significantly influenced by changes in BPD symptom severity (BSL-23 mean scores) or treatment response (responder, n=15 vs. non-responder, n=22).

## Discussion

This study aimed to investigate the neurobiological underpinnings of altered touch processing and its potential link to social dysfunctions in BPD. At baseline, BPD patients reported a significantly more negative attitude towards and significantly reduced comfort zones of touch for most social network members and body regions. Importantly, BPD patients showed decreased activation to CT-optimal vs. CT-suboptimal touch in the posterior insular cortex, which was significantly associated with more severe interpersonal problems. Thus, our findings point to a biological signature of the highly complex and debilitating social dysfunctions associated with BPD. Neither the behavioral ratings nor the neural responses significantly changed after four weeks of a residential DBT program despite significant improvement in overall BPD symptoms indicating trait-like disturbances in touch processing in BPD.

Our observation of a more negative attitude towards social touch in BPD patients is consistent with previous studies reporting that BPD patients exhibit lower needs for touch, less enjoyment of positive touch, and decreased importance of touch [26,28]. Reduced comfort zones of social touch in BPD patients were associated with more interpersonal problems, and not limited to a specific body region. This pattern was evident for all social network members except the romantic partner and female and male strangers. This could indicate that physical intimacy with romantic partners is less affected by pathological touch distortions, but given strong evidence that BPD patients engage in dysfunctional romantic relationships [46], a lack of significant difference for the romantic partner and female and male stranger might reflect ceiling or floor effects, respectively. Interestingly, touch allowance maps further indicate a reduced source effect in BPD patients, that is, BPD patients treat family members and friends more similarly to strangers than HC, as previously reported in other mental disorders regarding disgust in an interpersonal context [47]. By displaying higher social touch aversion across social network members and body regions, BPD patients may become depleted of physical and mental health-promoting benefits of interpersonal touch [48]. Previous studies have shown that interpersonal touch can reduce feelings of social exclusion [49] and loneliness [50], both central social impairments in BPD patients.

In the fMRI touch task, BPD patients showed a significantly reduced posterior insula response to CT-optimal touch which was associated with the severity of interpersonal problems in BPD patients. This pattern of results is clearly distinct from CT-unspecific changes observed in anorexia nervosa [51] or MDD [30] patients. The posterior insular cortex is a major projection region of CT fibers, transmitting information about affective properties of touch [22,23]. Consequently, it plays a central role in encoding emotional aspects of touch, not only by processing somatosensory features but also by integrating sensory perception and emotions [52]. Recent evidence suggests that BPD patients not only display an altered processing of pain [25] but also elevated somatosensitive thresholds [26,27]. It has been observed that reduced perceived intensity of pleasant touch is linked to higher BPD symptom load [26], while lower pleasantness of touch did not correlate with BPD symptom severity. Consistent with a recent study on affective touch in BPD [27], our findings revealed no CT-specific differences in touch comfort ratings in BPD compared to HC. Furthermore, we did not observe selective group differences for social relative to non-social touch, but this might reflect a partial social component in the non-social condition, as another human still delivered the brushes. Taken together, our results indicate that altered processing of affective somatosensory stimuli in BPD is not limited to the perception of pain stimuli [25], but also involves CT-optimal touch.

Importantly, the diminished activity within the posterior insula was associated with interpersonal problems, a cardinal symptom of BPD [7]. In MDD patients, higher social touch aversion partially mediated the relationship between depressive symptoms and interpersonal problems and this effect has been attributed to a negative impact on social communication [53]. Similarly, attenuated insular processing of CT-optimal touch could hinder everyday social communication in BPD, leading to more severe interpersonal problems. However, the relationship between depressive symptoms and interpersonal problems in MDD patients was mediated to a greater extent by an aversion to touch by less familiar people [53]. This further highlights the differences between MDD and BPD, as our results suggest a widespread negative attitude toward social touch that notably affects members of the inner social network. Interpersonal touch is crucial for the development of the social brain [54,55], and early experiences of social touch during infancy can significantly influence later attachment behavior [56]. We speculate that an aberrant processing of CT-optimal touch, potentially emerging early in social development, may have led to altered tuning of social brain networks, subsequently affecting interpersonal behavior later in life. Given that social dysfunctions are a complex multifactorial symptom of BPD, the dissociation of significantly improved interpersonal problems and persistent behavioral and neural disturbances in touch processing after the therapy could be explained by improvements in other domains like emotion regulation that also influence interpersonal relationships. The lack of a significant treatment effect, despite a reduction in symptom severity, suggests that pathological touch distortions are a trait-like aspect of BPD, similar to elevated pain and heat thresholds, which persist after 12 weeks of DBT [57].

However, future studies should test whether specifically targeting social touch processing in BPD can significantly reduce symptom burden and improve interpersonal functioning. Current DBT treatment approaches focus on mindfulness and interpersonal problems. Combining these elements, for instance, by teaching patients how to mindfully perceive and engage in touch, could be beneficial. A potential intervention might involve gradually introducing positive, controlled touch interactions with members of the patient’s social network, ensuring that clear boundaries are respected. This approach could not only help BPD patients to identify boundaries and achieve autonomy during touch experiences but also facilitate cognitive reappraisal of touch, which patients could then generalize to everyday situations. This idea aligns with the notion that traumatic experiences may lead to a negative bias in the processing of touch and altered cognitive appraisal of touch contexts [58]. Disrupted oxytocin signaling in BPD [59] may contribute to the impaired integration of sensory components. Consequently, addressing pathological distortions in touch processing through a combination of cognitive reappraisal strategies and oxytocin augmentation therapy may be a promising treatment approach for BPD.

Interestingly, the negative attitude towards and altered neural response to touch were not significantly moderated by trauma exposure in BPD patients. Similar to patients with Post-Traumatic Stress Disorder (PTSD), BPD patients report a negative bias toward social stimuli [60]. However, they display a distinct pattern of social dysfunctions [60]. While PTSD patients tend to exhibit increased automatic affective responses, BPD patients demonstrate a negative appraisal of stimuli, potentially due to impaired emotional regulatory control [12]. They not only display hypersensitivity to negative social cues but also a negative bias towards positive or neutral social stimuli. Consequently, they often exhibit severe mistrust, a heightened fear of rejection and abandonment, and difficulty distinguishing between social inclusion and exclusion [3]. They alternate between extremes of closeness and distance, as well as idealization and devaluation in their social relationships [3]. Finally, although traumatic experiences are highly common in BPD, it is important to note that not all patients diagnosed with BPD have experienced trauma [2]. The above-mentioned negative bias toward neutral or positive social cues might be intrinsic characteristics of BPD and not necessarily dependent on the severity of childhood trauma. Therefore, the attenuated processing of affective somatosensory stimuli in BPD may represent a learned self-protective mechanism resulting from at least subjectively perceived interpersonal challenges that increase the risk for future social dysfunctions. In line with a previous study [27] and in reference to Marsha Linehan’s statement on BPD patients having "no emotional skin" [1], we propose that BPD patients exhibit an impaired insula-mediated integration of sensory and emotional-motivational aspects of touch as a result of a learned psychological thickening of the skin to avoid being hurt by potentially harmful interactions, which might not be harmful to HC. However, further studies are needed to establish an empirical foundation for these speculations. We further did not detect significantly altered amygdala activation in response to touch, indicating that altered touch processing in BPD is not significantly related to a neural phenotype of threat hypervigilance [16,31]. Likewise, BPD patients exhibited no significantly altered activation in striatal regions in response to touch, which supports the notion of a BPD-specific neural touch signature not overlapping with MDD [30].

The present study has some limitations. We recruited a naturalistic cohort of BPD patients who presented with comorbidities and were under psychotropic medication. These sample characteristics may have contributed to the observed differences between BPD patients and HC. However, the present results are clearly distinct from a previous study involving MDD patients receiving antidepressants [30], which represented the most common class of psychotropic medication in our sample (see **Supplementary Tab. 2**). Furthermore, the true touch-related disturbances in BPD may even be underestimated in our study as the patients included had to be stable enough to undergo DBT and tolerate the experimental touch task. It is further conceivable that touch-related treatment changes would have become evident after the completion of all three DBT modules or longer follow-up assessments as improvements in interpersonal relations after DBT can occur time-delayed [61]. However, future studies should probe specific touch-focused interventions as we did not find significant changes in DBT responders compared to non-responders in the present sample. Furthermore, our study lacks a waiting-list patient group, which would help to disentangle time-dependent effects. Finally, to probe the disorder-specificity of the observed results, touch processing should be directly compared between BPD patients and those with other psychiatric conditions, such as MDD. Likewise, context effects should be tested by exploring differences in the neural response to touch from a familiar versus an unfamiliar person in BPD patients.

To conclude, this study shows that trait-like disturbances in touch processing and a disrupted insula-mediated integration of affective and sensory touch components may constitute a clinically relevant biological signature of interpersonal problems in BPD. As such, novel interventions targeting the altered processing of affective somatosensory stimuli may enhance long-term therapeutic outcomes by facilitating social functioning in patients with BPD.

## Statements

## Data Availability

All data produced in the present study are available upon reasonable request to the authors.

## Acknowledgment

The Center for Magnetic Resonance Research (CMRR) sequence was kindly provided by the University of Minnesota Center for Magnetic Resonance Research. Preprocessing of fMRI data was performed on the HPC Cluster CARL funded by the DFG under INST 184/157-1 FUGG. We thank Hannah Allmandinger, Paulina Piwkowski, Marlene Charlotte Holzhausen, Nick Michalek, Paul Grupe and Anja Sablotny for their help with data collection. Presented at the annual meeting of the Division of Biological Psychology and Neuropsychology of the German Psychological Society (DGPs) and the German Society for Psychophysiology and its Application (DPGA), Hamburg, May 29 – June 1, 2024.

## Statement of Ethics

The study was approved by the medical ethics committee of the Carl-von-Ossietzky University of Oldenburg, approval number 2020-101 and conducted in accordance with the latest revision of the Declaration of Helsinki. Participants provided written informed consent after receiving a complete description of the study.

## Funding Sources

D.S. was supported by a Research pool University of Oldenburg Medical Scientists grant (FP 2020- 047).

Author Contributions

J.V. and D.S. designed the experiment; J.V. conducted the experiments; J.V., D.P., and D.S. analyzed the data. J.V., D.S., D.P., R.H., and I.C. wrote the manuscript. All authors read and approved the manuscript in its current version.

Data availability

The data that support the findings of this study are not publicly available due to privacy reasons but are available from the corresponding author upon reasonable request and with Institutional Review Board approval.

