## Supplementary Information for "A neural signature of touch aversion and interpersonal problems in Borderline Personality Disorder"

### Supplementary methods

*Power-Analysis*

We used G*Power 3 [1] to conduct an a-priori power analysis for the study based on the effect size reported in a Cochrane Review on psychological therapies for patients with borderline personality disorder (BPD) [2]. The authors observed an overall medium effect size of $d$ = -0.6 for dialectical behavior therapy (DBT) treatment effects on borderline symptom severity. A different study [3] found the same effect size ($d$ = -0.6) for positive effects of DBT treatment on interpersonal problems in BPD. Because the study focuses on the effects of one DBT module (four weeks treatment), we assumed a reduced effect size of $d$ = 0.5. To replicate the abovementioned DBT treatment effect on borderline symptom severity and interpersonal problems in BPD (with $\alpha$ = 0.05 and power = 0.9, one-tailed paired-sample t-test) at least 36 patients have to be tested. Assuming a possible drop-out rate of 20-25% due to the longitudinal study design, we planned to test at least 50 patients.

Prior to study start, we conducted a pilot study with n=15 healthy controls (HC) to test our experiments. We used the data obtained during the social touch fMRI experiment to perform an fMRI-based power-analysis with NeuroPower [4]. To find significantly activated peak voxels for the contrast *social touch > non-social touch* (with $\alpha$ = 0.05 multiple comparison corrected using random field theory and power = 0.9, one-sample t-test) at least 18 HC have to be tested. As such, the planned sample size of n = 50 patients is sufficient to detect significant task effects. A previous study [5] observed large effect sizes for differences in social functioning between BPD patients and HC. Assuming a large effect ($d$ = 0.8) and considering an allocation ratio of 2:1, based on the assumption of a 50% treatment response rate in the patient group, (with $\alpha$ = 0.05 and power = 0.9, two-tailed two-sample t-test), at least 25 HC have to be tested. To account for possible drop-outs of 10-15%, we planned to test at least 30 HC.

*Study Design*

To further characterize BPD patients and to control for naturally occurring changes in the outcome measures, we included a control group involving healthy participants who did not receive an intervention. HC were recruited from the general population via online advertisements and flyers.

DBT at the Karl-Jaspers-Klinik comprises three modules, each followed by a break of varying duration. Patients were recruited from all three DBT modules (1$^{st}$ module n=23, 2$^{nd}$ module n=10. 3$^{rd}$ module n=4).

*Participants*

Axis I disorders were assessed by the Structured Clinical Interview for DSM-5 Disorders: clinician version (SCID-5-CV) [6]. HC additionally underwent the Zanarini Rating Scale for Borderline Personality Disorders (ZAN-BPD) [7] and completed the Assessment of DSM-IV Personality Disorders (ADP-IV) [8] to confirm the absence of any current psychological disorders. The average time between the last study appointment and admission to DBT was 4.15 ± 5.10 weeks. Accordingly, the HC group was assessed with a waiting period of five to nine week between pre- and post-measurement.

*Psychological and Clinical Assessments*

The Beck Depression Inventory-II (BDI-II) [9] and the Liebowitz Social Anxiety Scale (LSAS) [10] were used to measure depressive symptoms and social anxiety, respectively. A question based on a previous study [11] was administered to assess the frequency of interpersonal touch. Well-being before and after the fMRI touch paradigm was assessed on a visual analogue scale (VAS) from 0 to 100 (0 = ‘very bad’ to 100 = ‘very good’). Furthermore, inner tension was assessed on a VAS from 0 (not at all agitated) to 100 (very agitated) approximately half an hour before participants entered the MRI scanner. All participants completed questionnaires before and after four weeks of residential DBT/waiting interval. Depressive symptoms and social anxiety were only analyzed at baseline to characterize the sample. As part of a larger study, additional variables were assessed during residential treatment (reported elsewhere).

*Behavioral Data Analysis*

The statistical analyses were conducted using R [12]. Multilevel models with repeated measures were calculated using the nlme [13,14], lmerTest [15], and emmeans [16] packages of the statistical software program R. To assess possible group and treatment-related effects on well-being before and after the fMRI touch task, a mixed-design ANOVA with a between-subject factor group (HC, BPD) and a within-subject factor time (pre, post) was computed.

In cases where Levene's tests indicated heteroscedasticity, Welch's t-tests were applied. Additional Analyses of Variance (ANOVAs) were performed to investigate the effects of covariates for the pre-registered t-tests on questionnaire data and the TI.

Severity classification of BPD symptoms (BSL-23 scores) was performed according to [17].

*fMRI Touch Task*

For fMRI data, a T2*-weighted echoplanar (EPI) multiband sequence [18–20] was used to measure neural responses to touch (Repetition time (TR) = 850 ms, Echo Time (TE) = 30 ms, matrix size: 76 x 76, voxel size: 2.5 x 2.5 x 2.5 $mm^3$, slice thickness = 2.5 mm, distance factor = 0 %, field of view (FoV) = 192 x 192 $mm^2$, flip angle 62°, 48 slices). To control for inhomogeneities of the magnetic field, a fieldmap was obtained prior to the touch task and was included during preprocessing of the fMRI data (TR = 533 ms, TE (1) = 5.19, TE (2) = 7.65, matrix size: 64 x 64, voxel size: 3 x 3 x 3 $mm^3$, slice thickness = 3.0 mm, distance factor = 33 %, FoV = 192 x 192 $mm^2$, flip angle 60°, 35 slices). High-resolution T1-weighted structural images were collected at the same scanner (TR = 2000 ms, TE = 2.07 ms, matrix size: 320 x 320, voxel size: 0.8 x 0.8 x 0.8 $mm^3$, slice thickness = 0.75 mm, FoV = 240 x 240 $mm^2$, flip angle = 9°, 224 slices) and were included during the analysis of fMRI data. To optimize trial arrangements, the easy-optimize-x software (https://www.bobspunt.com/easy-optimize-x/) was used. Optimal designs were generated and randomly assigned to participants. Trial arrangements differed between pre- and post-measurement. Types and duration of touch were signaled via tones to the experimenter wearing headphones. The experimenter was trained to apply the tactile stimuli with a consistent speed and pressure. During the experiment, participants rated the comfort (German: Fürsorglichkeit) of each trial on a visual analogue scale (VAS) from 0 (not at all comforting, German: überhaupt nicht fürsorglich) to 10 (very comforting, German: sehr fürsorglich), including no touch trials. The participants completed two additional fMRI tasks (reported elsewhere).

*fMRI Data Analysis*

fMRI data were preprocessed using the standardized pipeline *fMRIPrep* 20.2.1 [21], which is based on *Nipype* 1.5.1 [22,23]. The first four volumes of each functional time series were discarded to allow for T1 equilibration. The pipeline included correction for magnetic field inhomogeneities and removal of movement components using Independent Component Analysis – Automatic Removal of Motion Artifacts (ICA-AROMA). Physiological noise was corrected using the Component Based Noise Correction Method (CompCor). The fMRI analysis was conducted using SPM12 (https://www.fil.ion.ucl.ac.uk/spm/), implemented in MATLAB release R2021a (The MathWorks, Natick, MA). The analysis involved a two-level approach, analyzing the data on single-subject and group level. General Linear Models (GLMs) were used, and GLM parameters were estimated using the restricted maximum likelihood method. The FAST model was applied to correct for autocorrelations. Hemodynamic responses to all five conditions (CT-optimal social touch, CT-optimal non-social touch, CT-suboptimal social touch, CT-suboptimal non-social touch, no touch) were modeled as boxcar functions with condition duration as length. The duration of ratings, the break between runs, three CompCor components explaining the most variance in the data, cosine regressors as a high-pass filter, and the mean global signal were included as nuisance regressors.

All ROIs were extracted using the WFU Pick Atlas Toolbox [24]. To define the ROIs, we used the Automated Anatomical Labeling (AAL) Atlas [25] for the bilateral amygdala and insula cortex, and the IBASPM 71 [26] for the striatum, which comprised the nucleus accumbens, caudate nucleus, and putamen. All ROIs were combined into a single ROI mask to adjust for multiple ROIs. Voxelwise habituation of the amygdala and striatum was investigated by calculating the ROI-specific mean response difference between the two runs (i.e. the two halves) of the touch paradigm. A second ROI-mask including only the amygdala and the abovementioned striatal areas was created to investigate habituation effects of touch vs. no-touch, CT-optimal vs. CT-suboptimal touch and social vs. non-social touch. A post hoc subdivision of the insula cortex into anterior and posterior subregions was performed according to [27] to better contextualize the significant finding within the framework of existing literature.

*fMRI Representational Similarity Analyses (RSAs)*

To explore group differences in neural patterns for the insula cortex and primary somatosensory cortex (SI) we performed ROI-based RSAs using the Decoding Toolbox (TDT) [28]. Similarities between all eight conditions (i.e., CT-optimal social touch run 1, CT-optimal social touch run 2, CT-optimal non-social touch run 1, CT-optimal non-social touch run 2, fast social touch run 1, CT-suboptimal social touch run 2, CT-suboptimal non-social touch run 1, CT-suboptimal non-social touch run 2) were calculated using Fisher z-transformed Pearson correlation coefficients and visualized in group-specific Representational Similarity Matrices (RSMs). Differences in mean correlation values were calculated for social vs. non-social touch and CT-optimal vs. CT-suboptimal touch for each group separately and were compared between groups using bootstrapped two-sample t-tests with 10000 replicates. The same procedure was applied to compare differences in neural patterns before and after four weeks of inpatient DBT within BPD patients using bootstrapped paired t-tests.

*Missing values*

Five BPD patients have missing data on years of education, and two BPD patients did not complete the Social Touch Questionnaire for the pre-measurement.

### Supplementary results

*Touch allowance maps*

The FDR-corrected z-thresholds with no correlation assumption varied from 2.84 to 4.08, depending on the number of participants identifying the respective social network member as part of their social network.

*Touchability Index (TI)*

To assess the influence of relationship type on the TI at baseline, we conducted a multilevel model with repeated measures. The model included the nine social network members (friend (f/m), stranger (f/m), partner, brother, sister, mother, father), group (HC, BPD) and the interaction of group and social network member as categorical fixed effect factors. The baseline model, including only the intercept, showed significant variance in intercepts across participants *(*SD: 0.13*,* 95% CI: 0.11,0.16*,* $X^2_{(1)}$=57.88*, p*<0.0001*)*. Different social network members (with female stranger as reference category), group (with HC as reference category), and various interactions of social network member and group predicted the TI (for parameter information, see **Supplementary Tab**. **4**).

To evaluate treatment-related effects, we employed a multilevel model with repeated measures, including the nine social network members, group, and time (pre, post) as categorical fixed effect factors. The analysis did not reveal a significant effect of time (with pre as reference category) or an interaction effect of time and group on the TI (all p-values > 0.18). The baseline model including only the intercept showed significant variance in intercepts across participants (SD: 0.13, 95% CI:0.11,0.16, $X^2_{(1)}$=167.70, *p*<0.0001), but slopes did not vary significantly across participants for the predictor time ($X^2_{(1)}$=0.35, *p*=0.84). For more information on model parameters, see **Supplementary Tab. 6**).

A third multilevel model with repeated measures evaluated treatment effects within BPD patients. The nine social network members were included as between-subject factor and time as within-subject factor. Results showed no significant main effect of time or interaction effect of time and social network member (all *p*-values > 0.23). The baseline model including only the intercept showed significant variance in intercepts across participants (SD: 0.17, 95% CI:0.16,0.18, $X^2_{(1)}$=35.35, *p*<0.0001), but slopes did not vary significantly across participants for the predictor time ($X^2_{(1)}$=0.95, *p*=0.62).

*Comfort Ratings*

The mixed-design ANOVA with group as between-subject factor (HC, BPD), presence of touch (touch, no touch) as within-subject factor, and comfort ratings of the fMRI task as dependent variable yielded a main effect of touch ($F_{(1,79)}$=79.87, $p$<.0001, $\eta_G^2$=0.40) such that touch stimuli were rated as more comforting than no touch stimuli. There was no main effect of group nor an interaction between group and touch (all $p$-values > 0.09).

*Well-being before and after the fMRI touch task*

A mixed-design ANOVA with group (HC, BPD) as between-subject factor and time (pre, post) as within-subject factor displayed no significant group or treatment-related effects (all $p$-values > 0.15).

*fMRI Touch Task Validation*

To validate that the fMRI touch task elicits activation in a touch-related brain network, we analyzed data of HC at baseline at both whole brain and ROI levels using a mask comprising the amygdala, insula cortex, and striatum. Touch compared to the no-touch control condition resulted in widespread activations on the whole-brain level, including the primary somatosensory cortex (SI), the putamen, and the insula cortex. The ROI analysis revealed significant activation within the bilateral insula cortex (right: peak MNI coordinates (x,y,z): 42, -10, 6; $t_{(27)}$=13.05; $p_{FWE}$< 0.0001, left: MNI: -40, -4, 0; $t_{(27)}$=11.78; $p_{FWE}$<0.0001), the bilateral putamen (left: MNI: -18, 14, 6; $t_{(27)}$=6.87; $p_{FWE}$=0.001, right: MNI: 20, 10, 4; $t_{(27)}$=6.24; $p_{FWE}$=0.004, MNI: 34, 4, 2; $t_{(27)}$=4.58; $p_{FWE}$=0.013) and the right amygdala (MNI: 22, -6, -14; $t_{(27)}$=5.85; $p_{FWE}$=0.010). A ROI analysis for CT-optimal touch compared to CT-suboptimal touch revealed significant activation within the right insula cortex (MNI: 40, -8, 4; $t_{(27)}$=7.26; $p_{FWE}$<0.0001) and the right amygdala (MNI: 24, 0, 14; $t_{(27)}$=5.21; $p_{FWE}$=0.040).

*Parameter Estimates correlation with Childhood Trauma or Symptom Severity*

There was no significant correlation between the parameter estimates of the significant peak voxel and BPD symptom severity (BSL-23 mean scores) or childhood trauma (CTQ total scores).

*Adding depression severity as a covariate*

In order to test whether the reduced processing of CT-optimal vs. CT-suboptimal touch in the posterior insular cortex may be confounded by depression severity, we added the BDI-II as a group mean centered covariate to the two-sample t-test comparing CT-suboptimal vs. CT-optimal touch in BPD patients compared to HC. The reduced processing within the posterior insular cortex remained significant even after controlling for depression severity (MNI: 40, -4, 2; $t_{(74)}$=4.62; $p_{FWE}$=0.027).

*fMRI Representational Similarity Analysis (RSA)*

We found no significant differences in similarity patterns for the insula cortex (all *p*-values > 0.46) and SI (all *p*-values >0.28) in BPD patients compared to HC. Additionally, there were no significant differences in neural patterns of the insula cortex (all *p*-values > 0.50) and SI (all *p*-values > 0.19) between pre and post-measurements within BPD patients.

*Changes in pre- and post-measurement compared between groups*

Mixed-design ANOVAs with group (HC, BPD) as between-subject factor and time (pre, post) as within-subject factor revealed no significant group x time interaction effects for social touch aversion (STQ scores, $F_{(1,66)}$=0.005, *p*=0.94) or comfort zones of social touch (TI scores, $F_{(1,66)}$=3.52, *p*=0.07). A mixed-design ANOVA investigating comfort ratings with group as between-subject factor (HC, BPD), time (pre, post) and presence of touch (touch, no-touch) as within-subject factors revealed no significant group x time interaction effects (all *p*-values > 0.73). Changes in neural responses towards touch versus no touch did not differ significantly between groups for the amygdala, striatum, or insula cortex.

*Moderation Effects of Childhood Trauma and Symptom Severity*

There were no significant moderation effects of symptom severity (BSL-23 mean scores) or childhood trauma (CTQ total scores) on social touch aversion (STQ total scores), comfort zones of social touch (total TI), comfort ratings of touch stimuli, and neural responses to touch vs. no-touch and CT-optimal vs. CT-suboptimal touch. We detected a significant CTQ x group interaction effect in the left caudate nucleus (MNI: -8, 16, -10; $t_{(73)}$=4.94; $p_{FWE}$=0.010) for touch versus no touch. The extracted parameter estimates of the significant peak voxel correlated significantly with the Childhood Trauma Questionnaire total score in HC ($r_{(79)}$=0.67, *p*<0.0001) but not in BPD patients ($r_{(79)}$=-0.10, *p*=0.512).

*Exploratory whole brain level analysis*

An exploratory whole brain level analysis revealed group differences in the habituation of the left middle cingulate gyrus for CT-optimal relative to CT-suboptimal touch (MNI: -2, -36, 36; $t_{(75)}$=4.86; $p_{FWE}$=0.045) prior to treatment. Habituation of the right cerebellum changed significantly after treatment within BPD patients for touch versus no touch (MNI: 2, -46, -48; $t_{26)}$=60.85; $p_{FWE}$<0.0001). Further, habituation of the right precuneus (MNI: 6, -64, 50; $t_{(59)}$=5.34; $p_{FWE}$=0.001) and the right middle frontal gyrus (MNI: 32, 18, 38; $t_{(59)}$=5.04; $p_{FWE}$=0.009) was altered in BPD patients after treatment compared to HC for touch versus no touch. Significance was assessed at cluster level with $p < 0.05$, family-wise error (FWE) corrected.

### Supplementary Figures

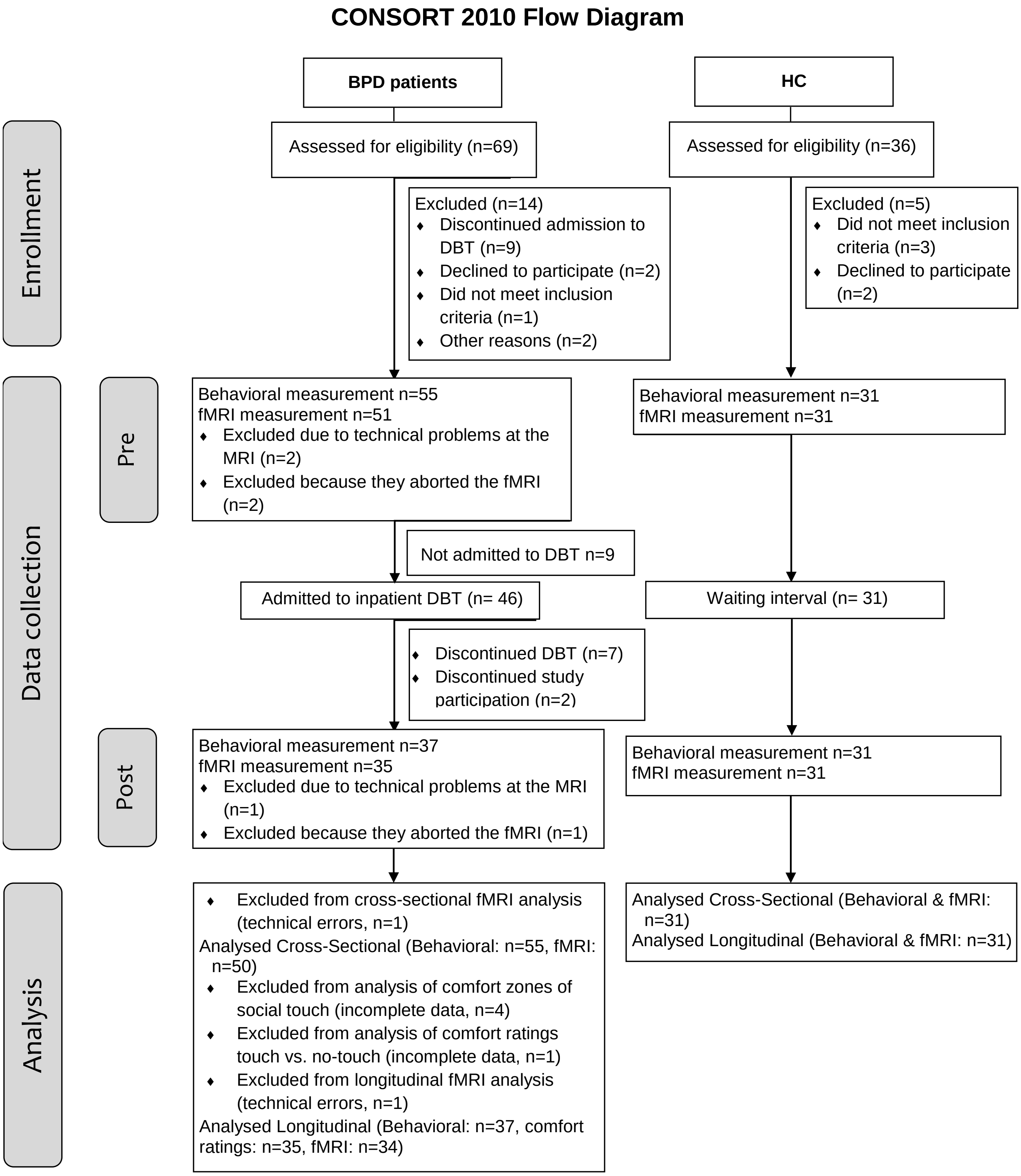


**Supplementary Figure S1:** CONSORT flow diagram of the entire study process.

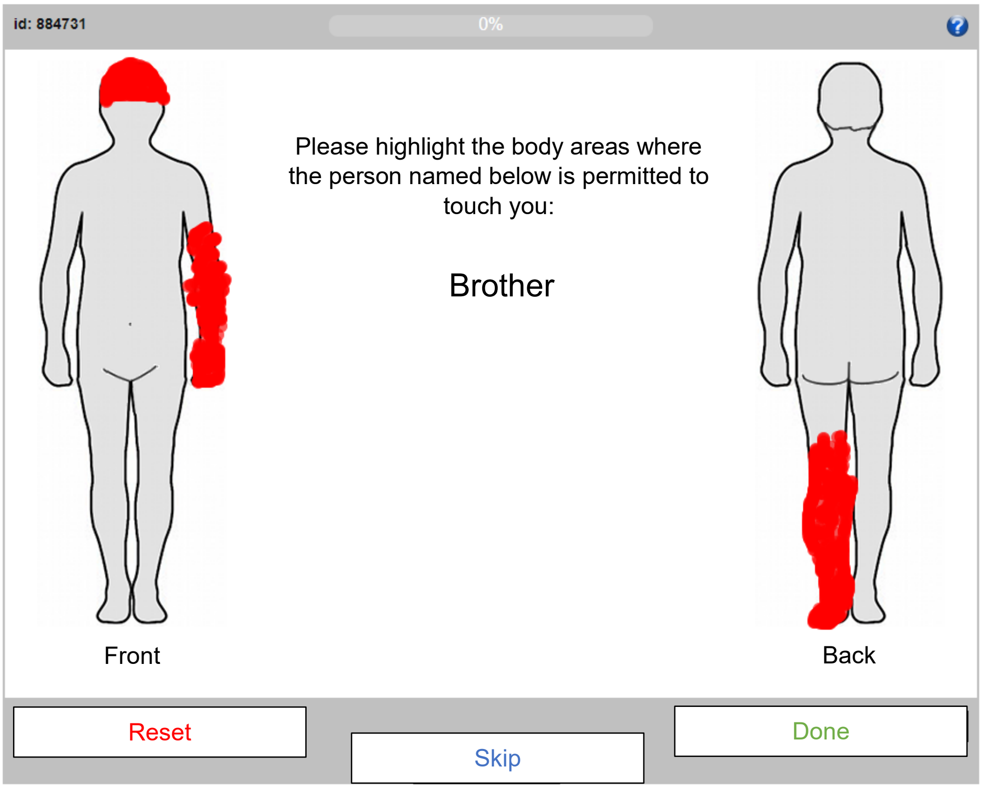


**Supplementary Figure S2:** Computerized task based on a previous study [29] to assess comfort zones of social touch. Participants were instructed to color body areas, where a specific social network member (presented in the middle of the screen) is allowed to touch them with their hand.

### Supplementary Tables

**Supplementary Table S1**: Sample description for patients with BPD and HC. The higher number of women than men reflects the distribution of BPD patients treated in clinical settings [30]. All study participants are Caucasian.

| | BPD patients (n=55) | | HC (n=31) | | |
|---|---|---|---|---|---|
| Characteristics | M ± SD or *n* | Min/Max | M ± SD or *n* | Min/Max | $X^2$ / *t-test, p* |
| *Demographic data* | | | | | |
| Age, years | 28.3 ± 10.2 | 18/57 | 28.0 ± 10.9 | 18/60 | 0.9 |
| Sex, male/female | 7/48 | | 4/27 | | 1 |
| Gender, male/female/diverse | 6/47/2 | | 4/27/0 | | |
| BMI | 29.6 ± 7.1 | 16.2/45 | 24.2 ± 4.5 | 18.8/36.6 | <0.0001 |
| Education, years | 13.6 ± 2.8* | 9/24 | 17.3 ± 3.0 | 13/24 | <0.0001 |
| *Psychological data* | | | | | |
| Depression (BDI-II) | 37.2 ± 12.5 | 6/57 | 3.6 ± 3.7 | 0/15 | <0.0001 |
| Social anxiety (LSAS) | 76.3 ± 30.6 | 1/124 | 24.5 ± 11.5 | 4/44 | <0.0001 |

*Notes. BDI-II, Beck Depression Inventory-II; BMI, Body Mass Index; BPD, borderline personality disorder; HC, healthy controls; LSAS, Liebowitz Social Anxiety Scale;M, Mean; n, number of available data points; SD, Standard Deviation; * data not available for 5 BPD patients*

**Supplementary table S2**: Axis I disorders, psychotropic medication, and frequency of interpersonal touch of BPD patients and HC.

| | BPD patients | HC |
|---|---|---|
| Characteristics | *n* | *n* |
| Axis I disorders (DSM-V criteria*)* | | |
| Any current comorbid diagnosis, yes/no | 48/7 | 0/31 |
| Any lifetime comorbid diagnosis, yes/no | 53/2 | 0/31 |
| *Psychotropic drugs* | | |
| Antidepressants | 35 | 0 |
| Antipsychotics | 22 | 0 |
| Psychostimulants | 6 | 0 |
| Anticonvulsants | 2 | 0 |
| *Interpersonal touch* | | |
| Less than once a week | 15 (27.3 %) | 3 (9.7 %) |
| More than once a week | 14 (25.5 %) | 8 (25.8 %) |
| More than five times per week | 13 (23.6 %) | 8 (25.8 %) |
| More than five times per day | 8 (14.5 %) | 8 (25.8 %) |
| More than ten times per day | 5 (9.1 %) | 4 (13.0 %) |

*Notes. n, number of available data points*

**Supplementary table S3**: Longitudinal sample description for patients with BPD and HC.

| Characteristics | BPD patients (n=37) | | HC (n=31) | | |
|---|---|---|---|---|---|
| | M ± SD or *n* | Min/Max | M ± SD or *n* | Min/Max | $X^2$ / *t-test, p* |
| *Demographic data* | | | | | |
| Age, years | 28.1 ± 10.7 | 19/57 | 28.0 ± 10.9 | 18/60 | 1 |
| Sex, male/female | 5/32 | | 4/27 | | 1 |
| Gender, male/female/diverse | 4/32/1 | | 4/27/0 | | |
| BMI | 29.7 ± 6.6 | 16.2/45 | 24.2 ± 4.5 | 18.8/36.6 | <0.001 |
| Education, years | 14.2 ± 2.7* | 10/24 | 17.3 ± 3.0 | 13/24 | <0.001 |
| *Psychological data* | | | | | |
| Depression (BDI-II) | 40.0 ± 9.4 | 6/57 | 3.6 ± 3.7 | 0/15 | <0.0001 |
| Social anxiety (LSAS) | 79.9 ± 29.6 | 1/124 | 24.5 ± 11.5 | 4/44 | <0.0001 |

*Notes. BMI, Body Mass Index; BPD, borderline personality disorder; HC, healthy controls; M, Mean; n, number of available data points; SD, Standard Deviation; * data not available for 2 BPD patients*

**Supplementary table S4**: Parameter information for the multilevel model at baseline.

| Predictor | β | SE β | 95% CI |
|---|---|---|---|
| Intercept | 0.55 | 0.04 | 0.47, 0.63 |
| BPD | -0.32 | 0.05 | -0.42, -0.21 |
| Friend (w) | >0.01 | 0.05 | -0.10, 0.10 |
| Friend (m) | -0.29 | 0.05 | -0.37, -0.20 |
| Partner | -0.39 | 0.04 | -0.48, -0.31 |
| Father | -0.05 | 0.04 | -0.13, 0.03 |
| Mother | 0.15 | 0.04 | 0.07, 0.24 |
| Brother | 0.17 | 0.04 | 0.08, 0.25 |
| Sister | 0.40 | 0.04 | 0.31, 0.48 |
| Stranger (m) | 0.17 | 0.05 | 0.07, 0.26 |
| Age | >0.01 | >0.01 | >-0.01, >-0.01 |
| Sex (m) | -0.04 | 0.05 | -0.14, 0.05 |
| BMI | >0.01 | >0.01 | >-0.01, >0.01 |
| BPD x friend (w) | 0.01 | 0.06 | -0.11, 0.14 |
| BPD x friend (m) | 0.21 | 0.06 | 0.10, 0.32 |
| BPD x partner | 0.22 | 0.06 | 0.11, 0.33 |
| BPD x father | 0.16 | 0.06 | 0.05, 0.27 |
| BPD x mother | 0.08 | 0.06 | -0.03, 0.19 |
| BPD x brother | 0.02 | 0.06 | -0.09, 0.13 |
| BPD x sister | 0.22 | 0.06 | 0.11, 0.33 |
| BPD x stranger (m) | 0.06 | 0.06 | -0.06, 0.18 |

*Note. SE, Standard Error, 95% CI, 95% Confidence Interval*

**Supplementary table S5**: Post-hoc pairwise comparisons of the TI.

| Social network member (contrast: HC - BPD) | EMM TI | | β | SE β | *p(cor)* |
|---|---|---|---|---|---|
| | HC | BPD | | | |
| Friend (f) | 0.70 | 0.46 | 0.24 | 0.05 | <0.0001 |
| Friend (m) | 0.50 | 0.33 | 0.16 | 0.05 | <0.01 |
| Partner | 0.95 | 0.85 | 0.10 | 0.05 | 0.12 |
| Father | 0.55 | 0.23 | 0.31 | 0.05 | <0.0001 |
| Mother | 0.72 | 0.41 | 0.30 | 0.05 | <0.0001 |
| Brother | 0.56 | 0.25 | 0.30 | 0.06 | <0.0001 |
| Sister | 0.72 | 0.46 | 0.26 | 0.06 | <0.001 |
| Stranger (m) | 0.16 | 0.06 | 0.10 | 0.05 | 0.12 |
| Stranger (f) | 0.26 | 0.15 | 0.11 | 0.05 | 0.11 |

*Note. EMM, estimated marginal means; SE, standard error. A Bonferroni-Holm correction was applied to adjust for multiple comparisons (p(cor)).*

**Supplementary table S6**: Parameter information for the longitudinal multilevel model.

| Predictor | β | SE β | 95% CI |
|---|---|---|---|
| Intercept | 0.56 | 0.04 | 0.48, 0.63 |
| BPD | -0.33 | 0.05 | -0.44, -0.22 |
| Post | 0.05 | 0.04 | -0.03, 0.14 |
| Friend (w) | >-0.01 | 0.05 | -0.10, 0.10 |
| Friend (m) | -0.29 | 0.04 | -0.37, -0.20 |
| Partner | -0.39 | 0.04 | -0.47, -0.31 |
| Father | -0.05 | 0.04 | -0.13, 0.03 |
| Mother | 0.15 | 0.04 | 0.07, 0.23 |
| Brother | 0.17 | 0.04 | 0.09, 0.25 |
| Sister | 0.40 | 0.05 | 0.31, 0.48 |
| Stranger (m) | 0.17 | 0.05 | 0.08, 0.26 |
| Age | >-0.01 | >0.01 | >-0.01, >-0.01 |
| Sex (m) | -0.03 | 0.05 | -0.13, 0.07 |
| BMI | > 0.01 | >0.01 | >-0.01, >0.01 |
| BPD x post | -0.06 | 0.06 | -0.17, 0.05 |
| BPD x friend (w) | > 0.01 | 0.07 | -0.12, 0.13 |
| BPD x friend (m) | 0.21 | 0.06 | 0.10, 0.13 |
| BPD x partner | 0.20 | 0.06 | 0.09, 0.30 |
| BPD x father | 0.16 | 0.06 | 0.05, 0.27 |
| BPD x mother | 0.09 | 0.06 | -0.02, 0.20 |
| BPD x brother | 0.03 | 0.06 | -0.08, 0.14 |
| BPD x sister | 0.27 | 0.06 | 0.16, 0.40 |
| BPD x stranger (m) | 0.96 | 0.06 | -0.04, 0.20 |
| Post x friend (w) | >0.01 | 0.07 | -0.13, 0.14 |
| Post x friend (m) | -0.05 | 0.06 | -0.17, 0.06 |
| Post x partner | -0.05 | 0.06 | -0.17, 0.06 |
| Post x father | -0.02 | 0.06 | -0.13, 0.10 |
| Post x mother | -0.06 | 0.06 | -0.17, 0.05 |
| Post x brother | 0.01 | 0.06 | -0.10, 0.13 |
| Post x sister | -0.06 | 0.06 | -0.16, 0.06 |
| Post x stranger (m) | -0.06 | 0.06 | -0.18, 0.07 |
| BPD x post x friend (w) | 0.02 | 0.09 | -0.16, 0.20 |
| BPD x friend (m) | 0.03 | 0.08 | -0.12, 0.19 |

| | | | |
|---|---|---|---|
| BPD x post x partner | 0.07 | 0.08 | -0.09, 0.23 |
| BPD x post x father | 0.05 | 0.08 | -0.11, 0.20 |
| BPD x post x mother | 0.08 | 0.08 | -0.07, 0.24 |
| BPD x post x brother | -0.03 | 0.08 | -0.19, 0.13 |
| BPD x post x sister | 0.04 | 0.08 | 0.11, 0.20 |
| BPD x post x stranger (m) | -0.05 | 0.09 | -0.22, 0.12 |

*Note. SE, Standard Error, 95% CI, 95% Confidence Interval*
